# SCORE: Serologic Evidence of COVID-19, Social, and Occupational Contacts in Healthcare Workers in a Sample of Long-Term Care and Acute Care Facilities in Southeastern Ontario (SCORE)

**DOI:** 10.1101/2024.05.03.24306818

**Authors:** Jorge L Martinez-Cajas, Beatriz Alvarado, Ann Jolly, Yanping Gong, Bradley Stoner, T. Hugh Guan

## Abstract

**Purpose:** Healthcare workers (HCW) have been an essential societal resource to face the COVID-19 pandemic. Early in the pandemic, they were at increased risk of contracting SARS-CoV-2 infection. We established a longitudinal cohort of HCW in an acute care hospital and four long-term care facilities in Ontario, Canada to follow the incidence of SARS-CoV-2 infection, the immune response to infection and/or vaccination, and the occupational, household and community factors related to their risk of infection.

**Participants:** Two hundred participants were recruited between November 2020 and July 2021. They completed a baseline survey, monthly surveillance data for 9-12 months, a post-Omicron-wave survey, and provided blood samples for anti-SARS-CoV2 antibody measurements. We collected data on host-related factors (humoral response to vaccines and SARS-CoV-2 infection) and environmental factors (social contact history, occupational, household and community conditions) to establish the main determinants of risk of SARS-CoV-2 infection.

**Findings:** Here, we describe the cohort demographics, occupational characteristics, SARS-CoV-2 vaccination status and COVID-19 infection risk during the cohort follow-up.

**Analyses:** The data from this cohort of HCW allows analyses on 1) the risk factors for SARS-CoV-2 infection, 2) the impact of the Omicron variant on the risk of infection; 3) the relationship between humoral responses and SARS-CoV-2 infection/vaccination and, 4) their relationship of SARS-CoV-2 infection and the community, household and healthcare facility-related exposures.

## Introduction

Healthcare workers (HCW) have been at the forefront of the response to COVID-19 worldwide and are an essential societal resource to face a pandemic. COVID-19 infections in HCW weaken the healthcare system’s capacity to respond to an epidemic by causing absenteeism. Before the arrival of effective vaccines, HCW fully relied on physical distancing, personal protective equipment (PPE) and institutional infection control protocols to avoid acquiring and limiting the spread of COVID-19. HCW continued to provide care to COVID-19 patients despite their concern about the effectiveness of available PPE (surgical masks in 2020) in preventing SARS-CoV2 infection [1]. In Canada, long-term care facilities (LTC) were disproportionately affected by COVID-19 during the first year of the epidemic. For instance, residents and HCW from LTC homes accounted for 16.6% and 16.9% of all cases Canada-wide, respectively [2]. The high risk of infection among LTC HCW during the early pandemic has been linked to multiple factors that include inadequate infection control practices while caring for COVID-19 patients,[3] a higher COVID-19 incidence rate in the region where the facility was located, older age of the buildings, a higher total number of beds, crowding,[4] a higher degree of interconnectedness in the facility and type of ownership (for-profit ownership being associated with higher risk compared with no-for-profit ownership)[5,6]. In the province of Ontario, the first two years of the COVID-19 epidemic presented substantial geographical variation in the incidence rates [7]. The Southeastern Ontario region remained a low-incidence area until the arrival of the variant of concern (VOC) Omicron wave offering an opportunity to explore the factors determining relative success in the epidemic control in this region with a focus on HCW. This is relevant as other respiratory viruses (Respiratory Syncytial Virus and Influenza) have since the winter of 2022 bounced back causing incidence peaks in the Northern hemisphere [8]. Between January and July 2020, HCW accounted for 19.4% of the SARS-CoV-2 infections in Canada but only 3% by June 2021 suggesting that the measures taken at community and institutional levels during this period effectively prevented the high rate of infection in HCW that had been seen in the early epidemic [9–11]. Even with the emergence of more infectious Alpha and Delta VOC, there were low incidence rates of COVID-19 in the Southeast region. Nevertheless, the arrival of the highly infectious Omicron wave in late 2021 cut through all barriers that had been previously effective, including high vaccination rates. Seroprevalence data in Canada have been published corroborating the lower effectiveness of the original COVID-19 vaccines to prevent transmission of the Omicron variants [10].

## Cohort objectives

In the fall of 2020, we established a prospective cohort of HCW to follow the evolution of the COVID-19 epidemic and to study factors that influence the risk of SARS-CoV2 infection. We proposed to study the risk in HCW adopting the framework of agent-host-environment triad as depicted in Figure 1. The agent SARS-CoV2 is a Betacoronavirus first discovered in China in December 2019 and caused a pandemic that spread rapidly around the world as it encountered an interconnected fully susceptible human population. SARS-CoV-2 infects host respiratory cells via interactions between its Spike (S) protein and the Angiotensin-Converting Enzyme ACE2 on the susceptible cell surface [12]. SARS-CoV-2 has a high mutational rate estimated at 1 x 10-3 substitutions per base (30nucleotides/genome) per year or 1 x 10-5 – 1 x 10-4 substitutions per base in each transmission event [13,14]which contributed to the emergence of a multitude of variants with enhanced transmissibility, differential virulence levels and immune evasion capabilities [15]. Surveillance of SARS-CoV-2 variants throughout the pandemic was performed in most countries and has allowed a detailed tracking of the evolution of this agent at a local and regional level. The human host population was naïve to SARS-CoV-2 and experienced a wide spectrum of infection severity from asymptomatic infection to severe systemic disease and fatal respiratory failure. The factors more strongly linked to higher disease severity are age, the presence of chronic conditions, compromised immune status and socioeconomic vulnerability. [16] [17] Later in the pandemic, vaccination status, vaccine type [18], and vaccine response [19,20] were found to deeply affect clinical outcomes of SARS-CoV-2 infections. The environment where hosts interact had a striking impact on how SARS-CoV-2 spreads. During the COVID-19 pandemic, it became clear that closer and longer social interactions, the absence of any form of personal protective equipment, and the characteristics of indoor facilities could promote the transmission of SARS-CoV-2. Therefore, in the present study, we gathered data that described to a feasible degree the factors encompassed by the agent-host-triad as felt pertinent to a sample of healthcare workers. In the case of occupation factors, studies support the role of type of institution, exposure to COVID-19 cases [21], social contacts [22], and type and use pattern of protective measures [23]; and as community factors, the role of community protective measures and social contacts influence the risk of COVID-19 acquisition in HCW [24] . The SCORE (***S****erologic Evidence of **CO**VID-19, Social, and Occupational Contacts in Healthcare Workers in a Sample of Long-Term Ca**re** and Acute Ca**re** Facilities in Southeastern Ontario) cohort* sought to 1) describe the prevalence and incidence of COVID-19 during the study period, based on self-report and serology; 2) describe the risk of COVID-19 from occupational and social contact patterns of HCW; and 3) describe the factors that affected the risk of acquiring COVID-19 in the cohort participants. In this paper, we describe the main exposures, outcomes, and serologic assays used to study this cohort in the epidemiological context of COVID-19 in Southeastern Ontario, Canada. This cohort recruited HCW of an acute care hospital and four LTC.

**Figure 1.**
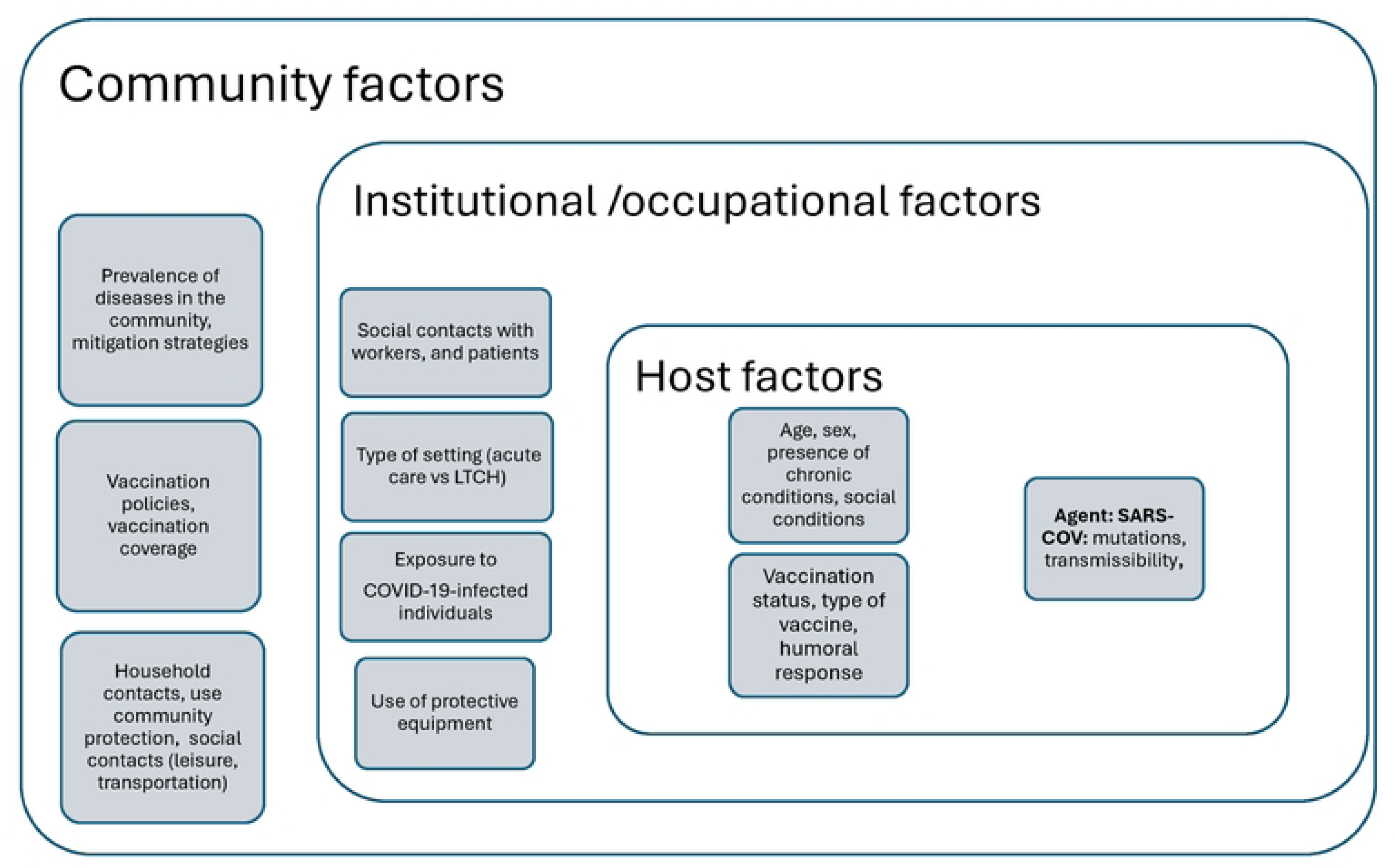
Framework to delineate risk factors for health care workers risk ofCOVID-19

## Epidemiological and Healthcare System Context

By the end of this cohort follow-up period (August 30, 2022), Ontario had experienced seven COVID-19 epidemic waves (S1 Table 1). During the first four waves, there was a distinct evolution of the epidemic with a higher incidence rate (5-10 times higher) in large urban centres versus a lower one in smaller urban and suburban areas [7,25]. This difference narrowed over time and virtually disappeared between the fifth and seventh waves predominantly caused by Omicron variants. Provincial mandates guided the healthcare response which presumably influenced the behaviour of HCW in the province, a sample of whom participated in this study. S2 Figure 1 summarizes the relevant provincial mandates and institutional regulations during the period covered by the SCORE cohort and relevant acute care hospital policies. Briefly, in January 2020 the first case of COVID-19 was confirmed in Ontario [26]. A few weeks later, the province declared a state of emergency with closures of most non-essential services [27]. The vast majority of cases of COVID-19 in HCW were laboratory-confirmed by PCR following provincial testing guidance [28,29]. Universal masking was instituted in April 2020 for all LTC homes in the province. In the acute care hospital, HCW were required to wear surgical masks and eye shields when involved in direct care of suspected/confirmed COVID-19 cases, N95 or higher level respirators during aerosol-generating procedures (AGP), with universal masking started in July 2020. N-95 respirators became the norm for direct care of COVID-19 cases in all healthcare facilities from wave five (Omicron wave) onward (Dec 2021). In the acute care hospital, screening for symptomatic individuals and high-risk contacts started as early as March 2020 and remained active during the study period (personal communication). In LTC homes, screening for symptomatic staff occurred daily province-wide and COVID-19 testing of asymptomatic staff was performed twice a week with PCR and/or antigen-based tests since May 2020 and throughout the cohort follow-up [30]. HCW vaccination started on Dec 15, 2020, to LTC staff in higher prevalence areas and was expanded later to regions of lower prevalence. In early March 2021, Ontario opted for a delayed second dose of the COVID-19 vaccine (four months rather than the manufacturer-recommended 21- or 28-day intervals) and focused on administering the first dose to as many high-risk people as possible [31]. Contact tracing capacity became overwhelmed during the Omicron wave in most jurisdictions in Ontario by the end of 2021. Between January and March 2022, Ontario gradually relaxed social gathering restrictions. By mid-March 2022, outdoor masking had become optional in the province [32].

**Table 1.** The number of patients and staff participants in the acute care hospital and long-term care homes included in the study.

| Institutional characteristics | **Acute Care** | **LTC home 1** | **LTC home 2** | **LTC home 3** | **LTC home 4/** |
| --- | --- | --- | --- | --- | --- |
| **Low vs high risk** |  | Low risk | High risk | High Risk | Low risk |
| **Size** | Large | Large/Urban | Medium/Urban | Large/rural | Large/Urban |
| **Number beds** | 440 | 243 | 60 | 253 | 174 |
| **Ownership** |  | Non-profit | For profit | Municipal | For profit |
| **Local incidence before September 2020** |  | Low (<150 cases per 100.000) | Low (<150 cases per 100.000) | Low (<150 cases per 100.000) | Low (<150 cases per 100.000) |
| **COVID unit** | Yes | No | No | No | No |
| **Recruitment period** | Nov 24th, 2020/ June 21 2021 | 18 Feb 2021/ 9 April 2021 | 29 December /29 April 2021 | 15 Jan 2021/19 Mar 2021 | 2 June 2021/2 July 2021 |
| **Number of participants recruited** | 142 | 9 | 19 | 27 | 11 |
| **Outbreaks since March 2020** | July 2021, Nov 2021 Jan 2022 Aug 2022 | April 2022 | **April 2020;** Nov 2020 | **May 2020;** April 2022 | Dec 2021, April 2022 |
| **Follow-up data** | 123 | 3 | 8 | 21 | 12 |
| **Follow-up data in August/September 2022** | 112 | 3 | 7 | 21 | 8 |

Source: Ontario LTCH database, analysis done by authors

### Ethics approval

This study was approved by the Queen’s Ethics Board for Research in Humans; ethics reference: DEMD-2405-20. All participants provided written (electronic) consent. The consent form covered information on (i) the content and duration of the questionnaire; (ii) the use of blood samples; (iii) the need for monthly surveillance; and (iv) the need for potential further follow-up contact. A second consent was obtained from participants to collect data during the Omicron wave which went beyond the originally planned 9-12 month follow-up period. No personal, identifying information was collected through the study questionnaires.

## Establishing the Cohort

The SCORE cohort sought to explore the evolution of the COVID-19 pandemic in a suburban region of Ontario, Canada. It started with the recruitment of HCW in an acute care hospital focusing on those more likely to interact with patients with either suspected or confirmed COVID-19: the COVID-19-dedicated unit, the emergency department (ED) and the intensive care unit (ICU), which were deemed the high-risk settings. Subsequently, we disseminated invitations for all HCWs in the hospital through posters and departmental emails. Additionally, we invited ten LTC homes to participate, from which four agreed, two had experienced COVID-19 outbreaks before recruitment and two had not. The two LTC homes that experienced previous COVID-19 outbreaks were categorized as high-risk settings while the two that had not were categorized as low-risk settings. In the participating LTC homes, HCWs were invited initially with the assistance of managers who forwarded email invitations, and through posters that contained a QR code directing potential participants to contact the study coordinator. Individuals were eligible to participate in the study if they were HCW in any of the participating institutions, were able to provide consent, and were willing to remain engaged during the 9-12-month follow-up. The surveys were completed online except in one LTC home where HCW had inconsistent access to institutional e-mail and required paper-based surveys. A sample size of 200 HCW provided sufficient power to find statistically significant differences between the prevalence of seropositivity or self-reported COVID-19 of 15% in settings with previous outbreaks or HCW in high-risk areas in the acute care hospital (COVID unit, intensive care units, emergency room) compared with a seroprevalence or COVID-19 positivity of 3% in settings with no previous outbreaks or HCW in low-risk areas in the acute care hospital. Table 1 describes the five institutions included in the study. Notably, no institutional LTC outbreaks of COVID-19 occurred in the participating LTC homes since the start of recruitment until November 2021. The first COVID-19 outbreak in the acute care hospital occurred in July 2021.

The cohort recruited 206 HCW who agreed to participate; 200 of whom provided complete baseline data, 168 (81%) provided two data points between May 2020 and July 2022, and 150 (72%) provided an additional (third) data point between August and September 2022 (Figure 2).

**Figure 2.**
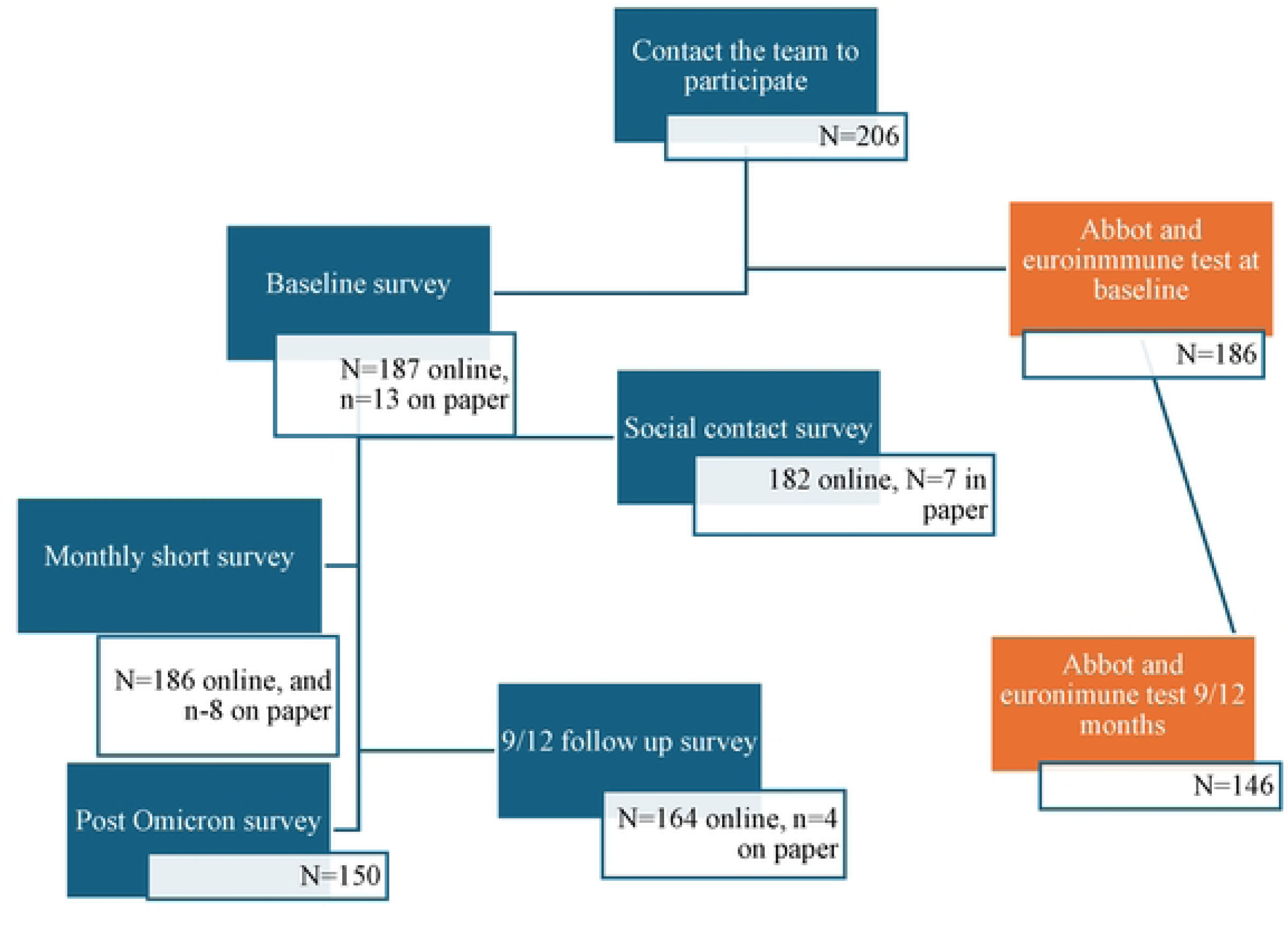
Recruitment and specimen collection of the SCORE cohort

Sixty-eight percent of the sample originated from the acute care centre. The majority of respondents were women, (85%), with an average age of 37 years. Nearly 60% of the sample were nurses, nurse assistants or personal support workers (PSW) (Table 2). Thirty-six percent had at least one chronic condition and 10% smoked tobacco. Most, (90%) were born in Canada, and 14% self-identified as a visible minority (based on religion, race, or gender). Differences in demographic characteristics were observed between HCW in the acute care hospital vs the LTC homes, with the former being younger, more economically self-sufficient, having a lower smoking rate and fewer chronic conditions (Table 2).

**Table 2.**
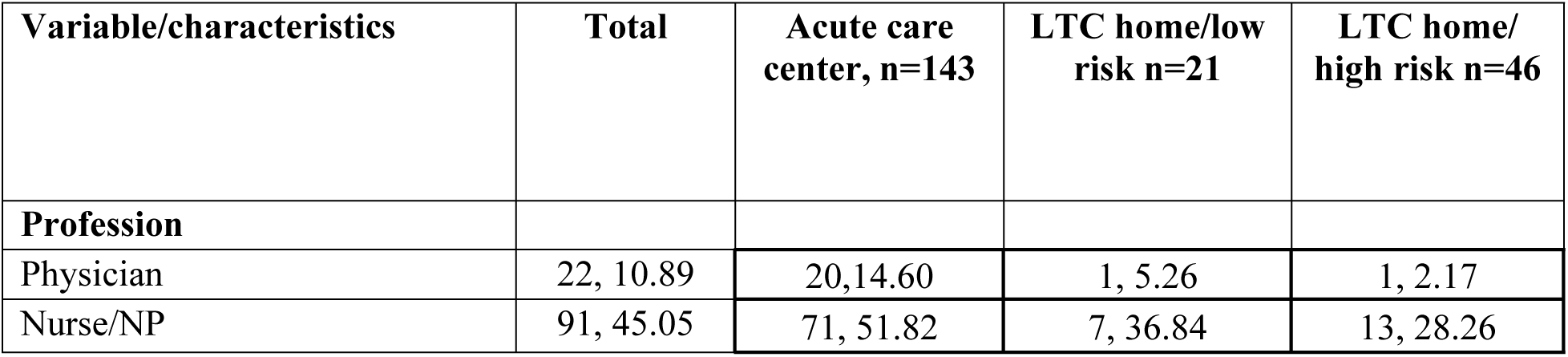
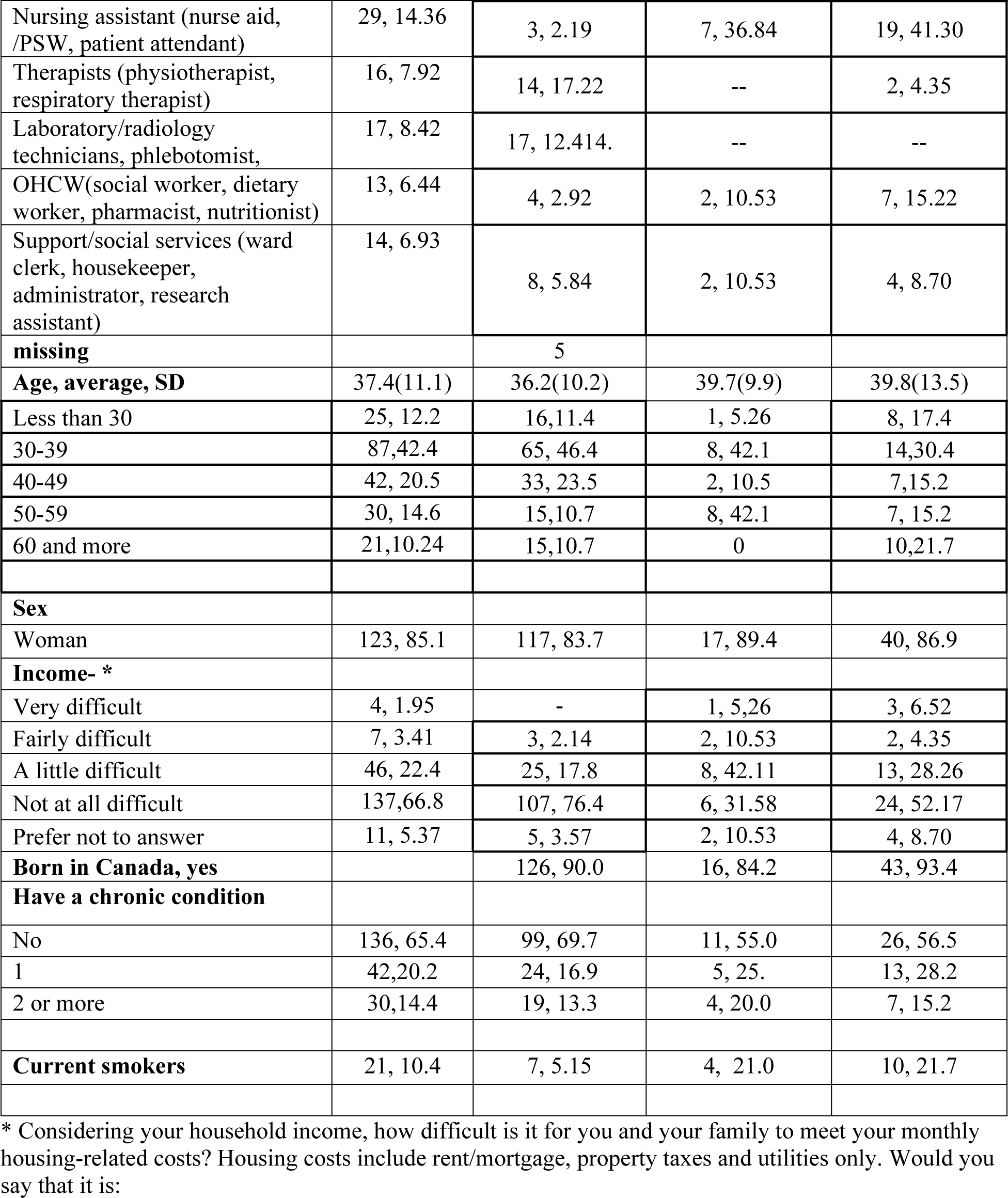
Descriptive data, sociodemographic characteristics of the participants in the SCORE, total and by settings.

The cohort retention rate was 72%. The main reasons for the loss of follow-up were relocating to another city and changing work site. Participants who were lost of follow-up were more likely to be nurse assistants or personal support workers (PSW), reported less sufficient income, and were more likely to be smokers. There was more participant loss in the LTC homes than in the acute care institution (S1 Table 2).

### Cohort follow-up

The SCORE cohort was initially designed to include a baseline assessment (questionnaire and blood draw), monthly questionnaires on infection, exposure and vaccination status, and a second questionnaire and blood draw 9-12 months after the initial assessment (Figure 3). The follow-up covered waves 1-7 of the COVID-19 epidemic in Canada (S1 Table 1 and Figure 3). Enrollment in the cohort started in November 2020 and the baseline was completed in June 2021. Monthly surveys started in December 2020 and ended in July 2022. Two blood samples were collected over the study period, at the beginning of the study between January and August of 2021 (the baseline or T1 sample) and then 9-12 months after, between October of 2021 and August of 2022 (T2 sample). The mean time between the two blood specimens was 48 weeks (range 36-80 weeks). All blood samples were collected after completion of the questionnaires, with a delay between the initial questionnaire and the first blood sample of 26 days (SD: 25) and between the 9-12 month questionnaire and the second blood sample of 48 days (SD: 43). Upon the arrival of the Omicron wave, we applied an additional survey to gather data on the impact of this wave.

**Figure 3.**
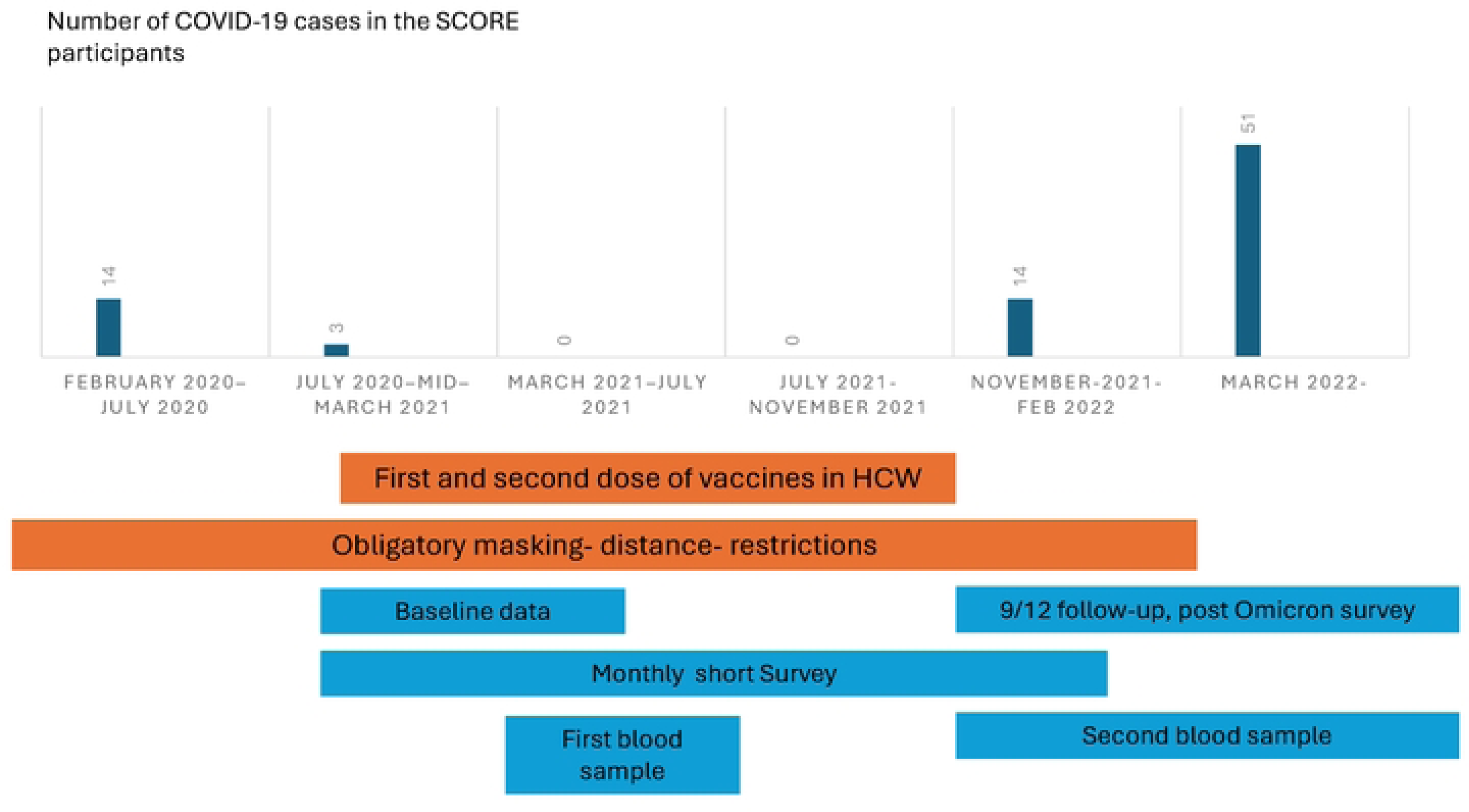
Number of cases and collection of data over the study period categorized by COVID-19 waves in **Canada**

### Measurements

We collected self-reported sociodemographic, occupational, household, and social contact data using the baseline questionnaire, a social contact questionnaire, contact tracing analysis of public health records, two follow-up surveys and monthly surveillance (Table 3).

**Table 3.**
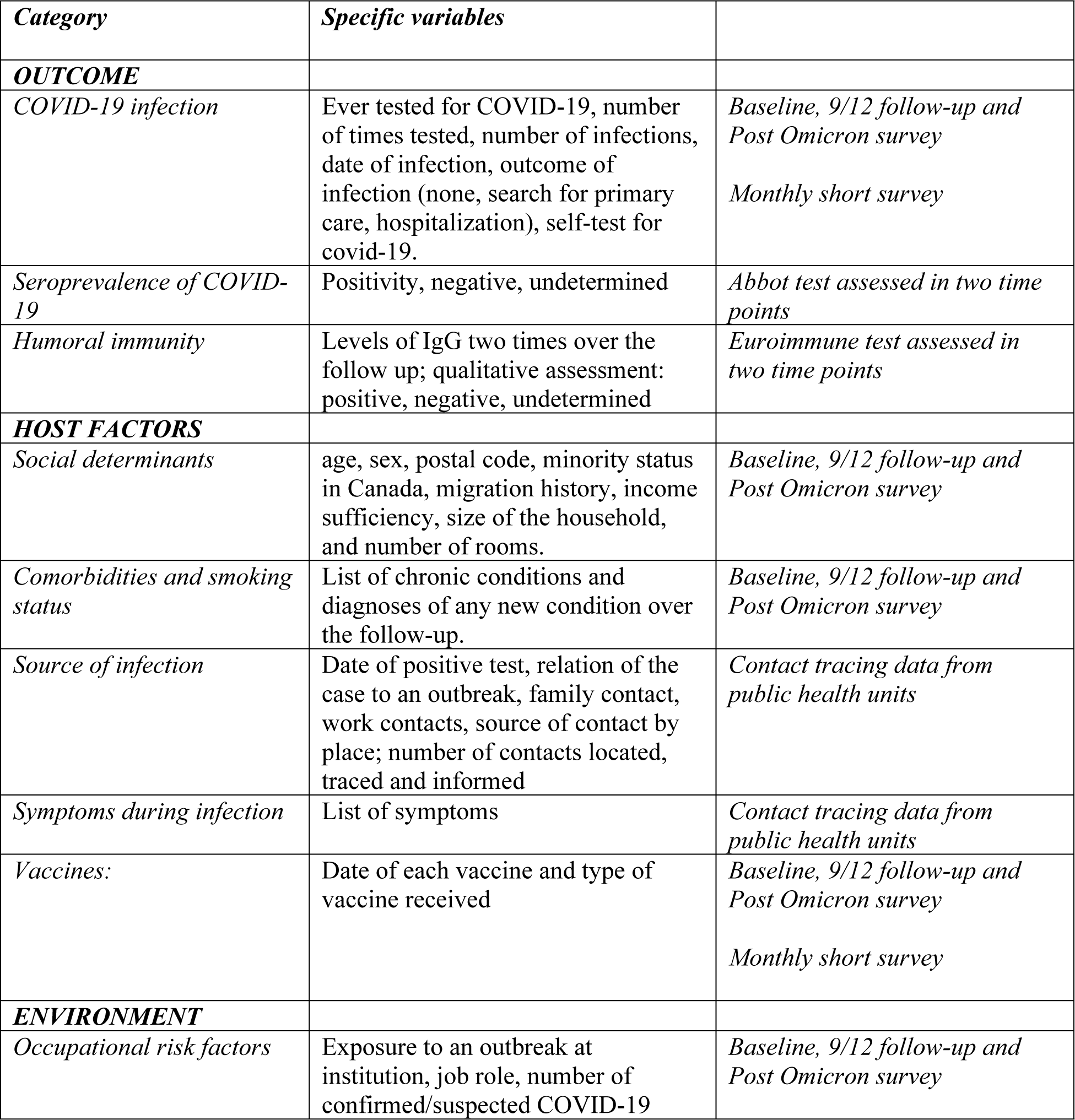
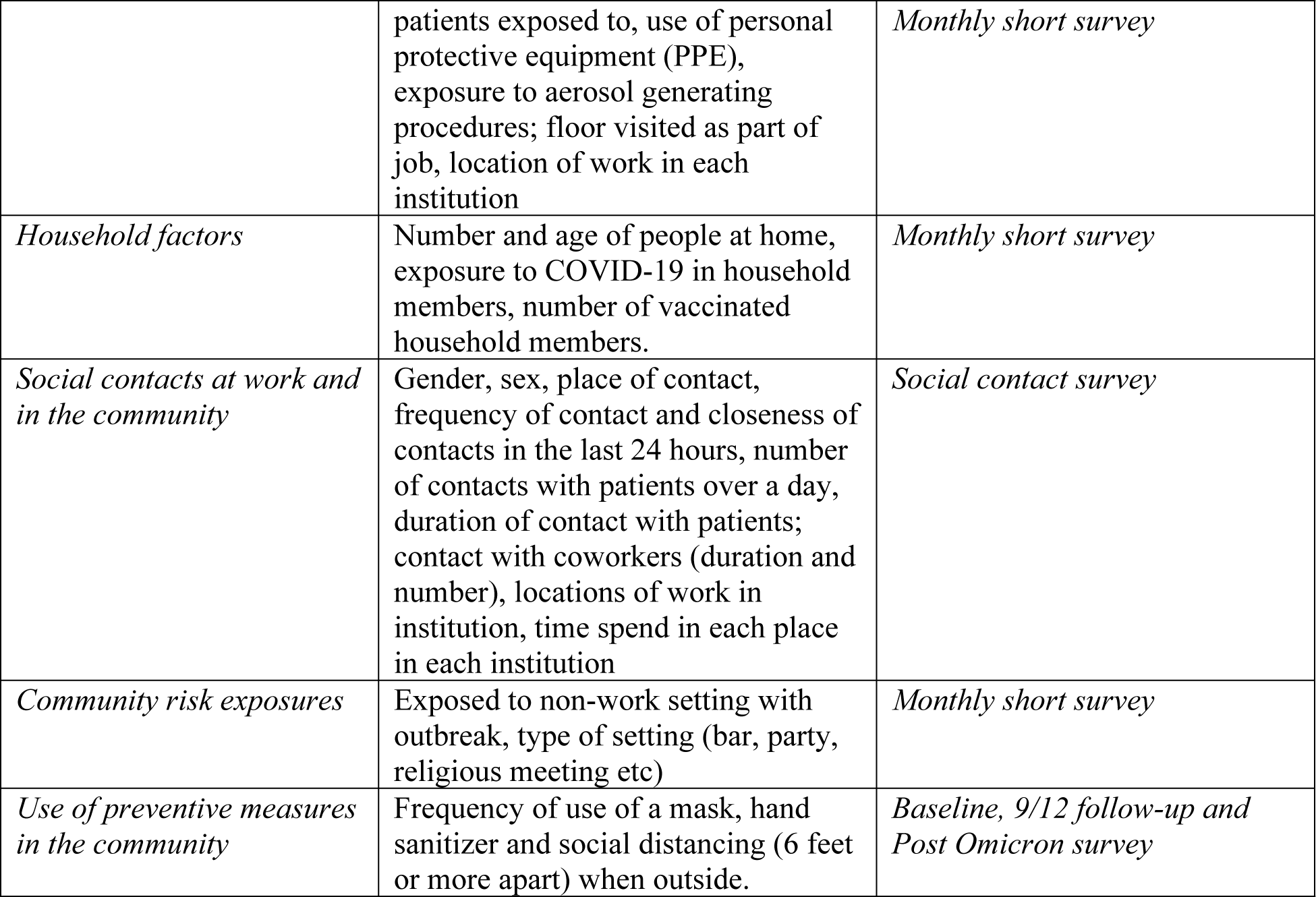
Variables and sources in the SCORE study.

**Our primary outcome was SARS-CoV-2 infection**, determined by the self-reported positive test for COVID-19. All LTC HCW in the province of Ontario were tested for SARS-CoV 2 infection using PCR testing when symptomatic or screened weekly or biweekly with either PCR or antigen-based tests when asymptomatic (between May 2020 and June 2023) [33]. HCW at the acute care institution were required to undergo PCR-based COVID-19 testing when symptomatic or when suspected to have had close contact with a COVID-19 case (Infection Control office of ACH, personal communication). Participants recorded the exact date of the positive test which was used to define the outcome. Importantly, HCW could access their test results and COVID-19 test results and vaccination history via the MOH COVID-19 registry.

**Factors related to the risk in HCW in each setting** were selected based on the available literature and our framework (Figure 1). These include 1) <u>host-related factors</u>, such as sociodemographic variables, health-related variables, variables related to COVID-19 vaccines, and the humoral response to vaccines. 2) <u>occupational variables</u>, including setting, use of protective measures, exposure to COVID-19 cases, exposure to AGP, and number of contacts at work. In the case of setting, it was further classified as i) high-(ICU, COVID unit, emergency room) and low-risk (other hospital units) acute care settings; ii) LTC HCW who worked at facilities that had COVID-19 outbreaks (higher risk LTC homes) and those who worked in LTC homes that had not had COVID-19 outbreaks (lower risk LTC) before the study onset. 3) <u>Community related factors</u> included household size, contacts at home, leisure school and transportation activities, and use of protective measures in the community. Questionnaires are available upon request to the corresponding author.

#### Assessment of humoral response

We used two antibody tests to detect previous infection with SARS-CoV-2 [34–36]. The first test was the Abbott SARS-CoV-2 IgG assay (Abbott Diagnostics, Abbott Park, Illinois, United States) which detects IgG antibodies against the Nucleocapsid (N) protein of the SARS-CoV-2 virus. For this test, a relative light unit index (S/C) with a result of > 1.4 was considered positive for past COVID-19 infection. This assay performs with modest sensitivity (approx. 70%) but high specificity ≥ 90% [37]. The second test was the semiquantitative Euroimmun Anti-SARS-CoV-2 ELISA IgG (Medizinische Labor diagnostika AG, Lübeck, Germany) which detects antibodies against SARS-CoV-2 S1 domain of the spike protein including the immunologically relevant receptor binding domain (RBD). This assay offers three possible interpretations: negative if the ratio is <0.8, borderline if the ratio is ≥ 0.8 to 1.1 and positive if the ratio is ≥ 1.1. The latter assay was used during the first phase of recruitment but abandoned when vaccination started because of cross-reactivity with vaccination-induced antibodies and because it did not allow quantification of high antibody levels expected to be induced by vaccination. A third ELISA test was used to quantify IgG antibodies against the SARS-CoV-2 spike receptor–binding domain (EUROIMMUN, product number: EI 2606-9601-10) [38]. This quantitative method offers a linear range between 3.2 to 384 BAU/mL (binding antibody unit). Samples with results over 384 BAU/mL were diluted by a factor of 20 to 30-fold to obtain numeric results. A cut-off of 35.2 BAU/mL was used to determine the seroconversion as recommended by the manufacturer [34].

## Results to date

### Prevalence and incidence over time

In the context of the COVID-19 epidemic in Ontario, the SCORE cohort reflects the evolution of the epidemic in Southeastern Ontario. S2 Figure 2-4 describe the cases since the start of the epidemic in the three regions where the participating institutions are located. COVID-19 cases in the SCORE cohort mimic the distributions in the region (Figure 4). There were relatively high number of cases at the beginning of the epidemic, with 16 of 206 (7.6%) HCW reporting having had COVID-19 between March and July 2020. During the monthly follow-up, only three participants reported incident COVID-19 cases until January 2022. New diagnoses started to be reported in January 2022 with a peak in cases between March and June 2022. A total of four (n=5) reinfections were reported or identified in the SCORE cohort. Two participants (no 2 and no 3) had both of their infections before Omicron wave (S1 Table 3), two participants had one infection pre-Omicron and another during the Omicron waves (Participants 1 and 5) and one participant reported two infections during the Omicron wave.

**Figure 4.**
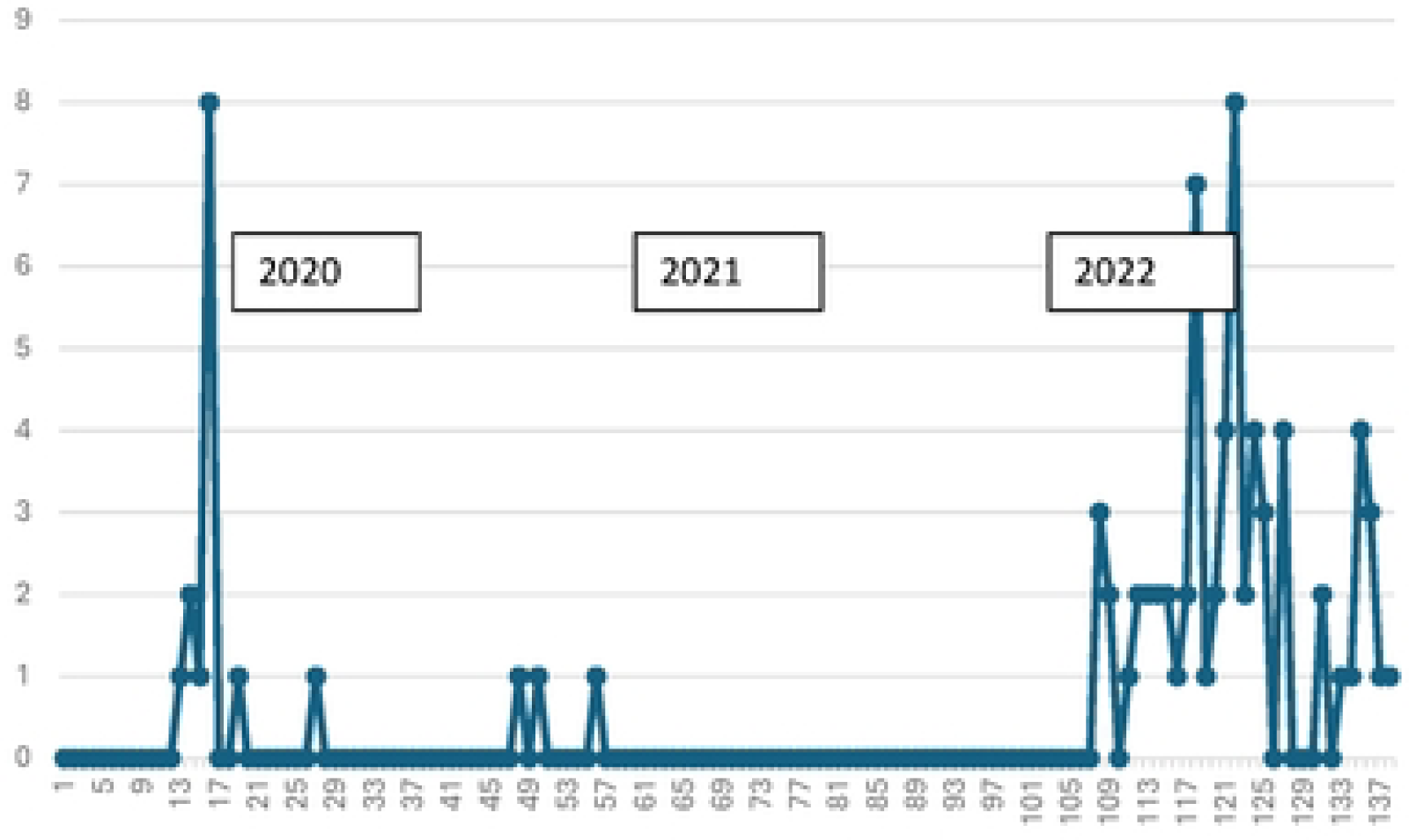
Distribution of cases over time in the SCORE participants

HCW from the acute care institution had a higher risk of infection during the Omicron wave compared to preceding waves (OR=7.64 CI95%: 4.24-13.7); while in the LTC homes at high risk, HCW experienced a similar risk of infection before and during the Omicron wave (OR: 1.76; CI95%: 0.63-4.9). For those in LTC homes at low risk the odds ratio was not calculated as there were no cases before Omicron (Table 4), and new cases occurred only later between (March and July 2022). Similarly, in HCW who had worked in high-risk settings in the acute care hospital, the risk of COVID-19 was observed at the beginning of March 2022 (Table 5). The excess risk of COVID-19 at the beginning of the epidemic in LTC homes HCW in Canada is well known and is consistent with the high burden of the epidemic in those settings [39].

**Table 4.** Estimated hazards using survival discrete analysis expressed as percentages and odds ratios in the whole sample and by setting.

| **Interval in months** | **Hazard %(logit OR)**<br>**Whole sample** | **Hazard %(logit OR)**<br>**Acute care sample** | **Hazard %(logit OR)**<br>**LTC no outbreak/Low risk** | **Hazard %(logit OR)**<br>**LTC outbreak/high risk** |
| --- | --- | --- | --- | --- |
| Feb 2020 - Jun 2020 | 6.25 (ref) | 1.41 (ref) | 0 | 23.9 (ref) |
| Jul 2020 - Nov 2020 | 1.03 (OR=0.15)* | 1.43 (OR=1.01) | 0 | 0 |
| Dec 2020 - April 2021 | 1.04 (OR=0.16)* | 1.45(OR=1.02) | 0 | 0 |
| May 2021 - Sep 2021 | 0 | 0 | 0 | 0 |
| October 2021/Feb 2022 | 8.23 (OR=1.34) | 7.56 (OR=5.7)* | 0 | 14.3 (OR=0.53) |
| March 2022/July 2022 | 37.31 (OR=8.9)* | 42.0 (OR=50.7)* | 18.2 | 26.1 (OR=1.12) |
| August /Sept2022 | 1.23 (OR=0.18) | 1.75 (OR=1.25) | 0 | 0 |

**Table 5.** Hazards calculated using survival discrete analysis expressed as percentages and odds ratios in the Acute care center.

| **Interval in months** | **Hazard %(logit OR)**<br>**Acute care sample/ high risk** | **Hazard %(logit OR)**<br>**Acute care sample/ low risk** |
| --- | --- | --- |
| Feb 2020 - Jun 2020 | 0 | 1.98 (ref) |
| Jul 2020 - Nov 2020 | 2.44 (ref) | 1.01 (0.50) |
| Dec 2020 - April 2021 | 0 | 2.04 (1.03) |
| May 2021 - Sep 2021 | 0 | 0- |
| October 2021/Feb 2022 | 0 | 10.3(5.7)* |
| March 2022/July 2022 | 37.9 (24.4)* | 43.6 (38.3)* |
| August /Sept2022 | 0 | 2.56 (1.30) |

Regional genotype surveillance depicts the transition of SARS-CoV 2 variants since the beginning of the epidemic (S2 figure 5). The distribution of the predominant sublineages in the three regions corresponds to the ones reported in the province of Ontario, where sublineages were identified as predominant BA.1/BA.2 between January 2−June 18, 2022, and BA.4/BA.5 between June 19−November 26, 2022. Assuming infections in this cohort by the reported date, they resemble the pattern seen in Ontario. Overall, 17% of the cases were non-Omicron lineages, 68.4% occurred during BA.1/BA2 predominance and 14.4% when BA4/BA5 was predominant

**Figure 5.**
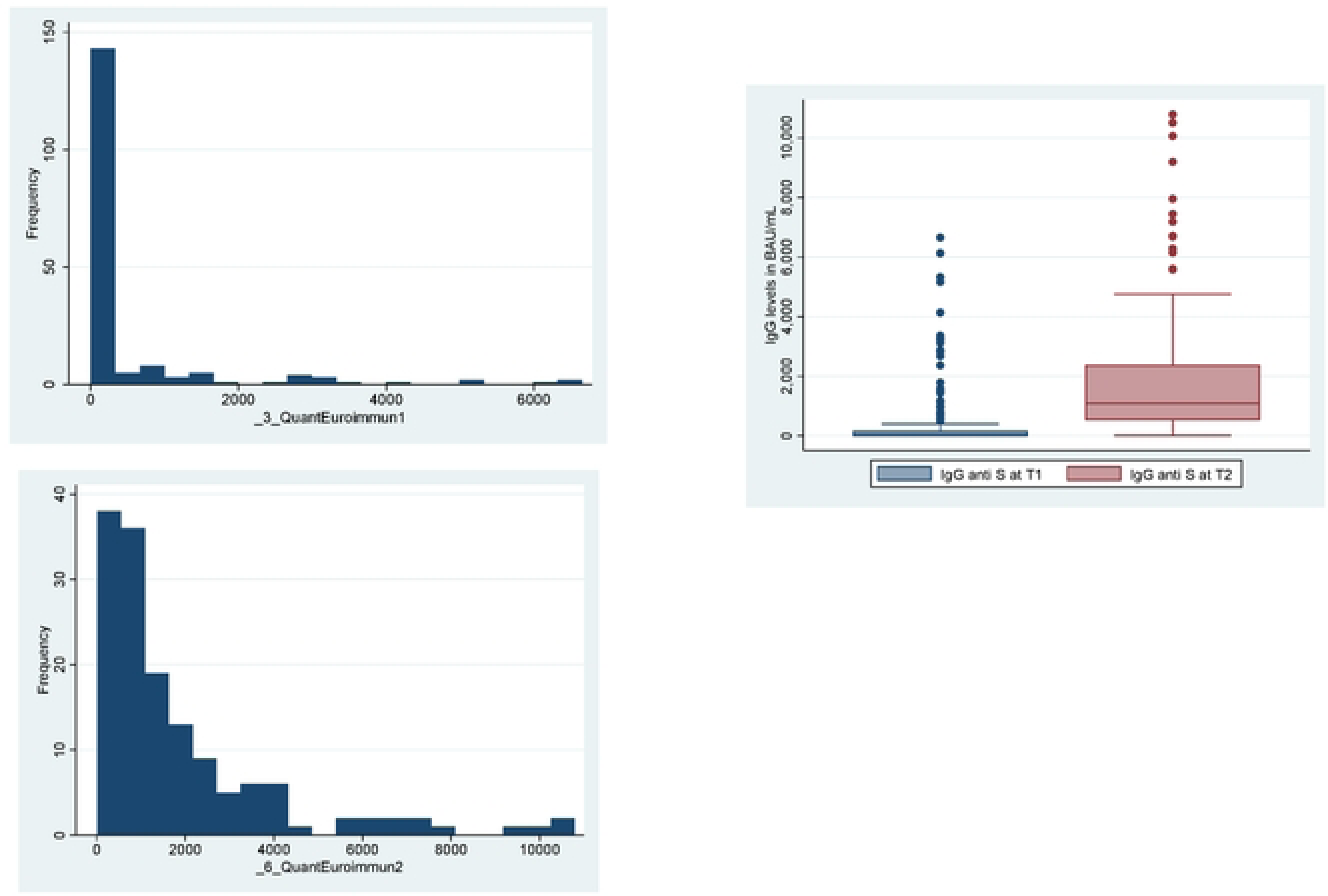
Histogram and boxplot dcpic1ing the distribution oflgG ofan1iS in participants of SCORE a1Tl and T2

### Risk of occupational exposure over time

The risk of exposure to COVID-19 varied throughout the cohort follow-up (Table 6). At baseline, 50% of the participants had provided care to a COVID-19 case, which rose to 67.9% during the Omicron wave. Nearly 18% of HCW reported caring for more than 20 COVID-19 patients at baseline, which rose to 38% during the Omicron wave. At baseline, 8% of the cohort had worked in a COVID-19 unit, 12% in the emergency room, and 20% in the ICU. During the Omicron wave, 11% worked in a COVID-19 unit, 24% in the emergency room, and 34% in the ICU. Interestingly, and in concordance with institutional recommendations, a higher proportion of HCW (94%) used N95 respirators during the Omicron wave than earlier in the epidemic (47%), while the use of surgical masks was reduced when providing care to COVID-19 patients (S2 Figure 6), and was replaced by the use of N95 or higher respirators. At the beginning of the epidemic, some HCW from the acute hospital were already using N95 respirators, though this was not the case for HCW in the LTC homes. This trend changed over time so that 80% of HCW used N95 when caring for COVID-19 patients/residents (S2 Figure 7).

**Table 6.**
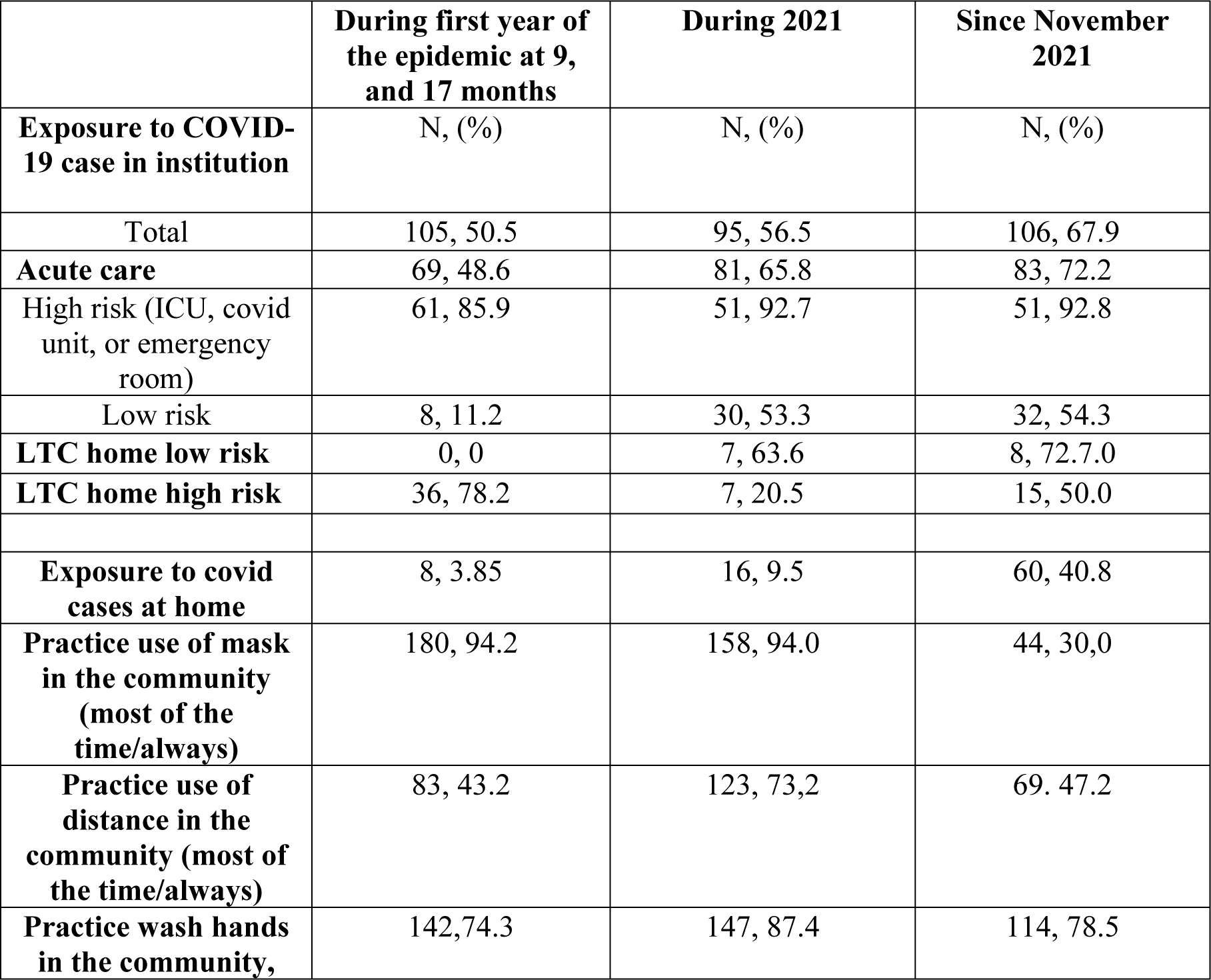
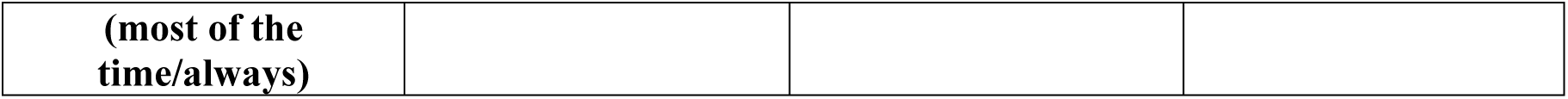
Description of the cohort in terms of risk exposure to COVID-19. Occupational and community exposures.

**Figure 6.**
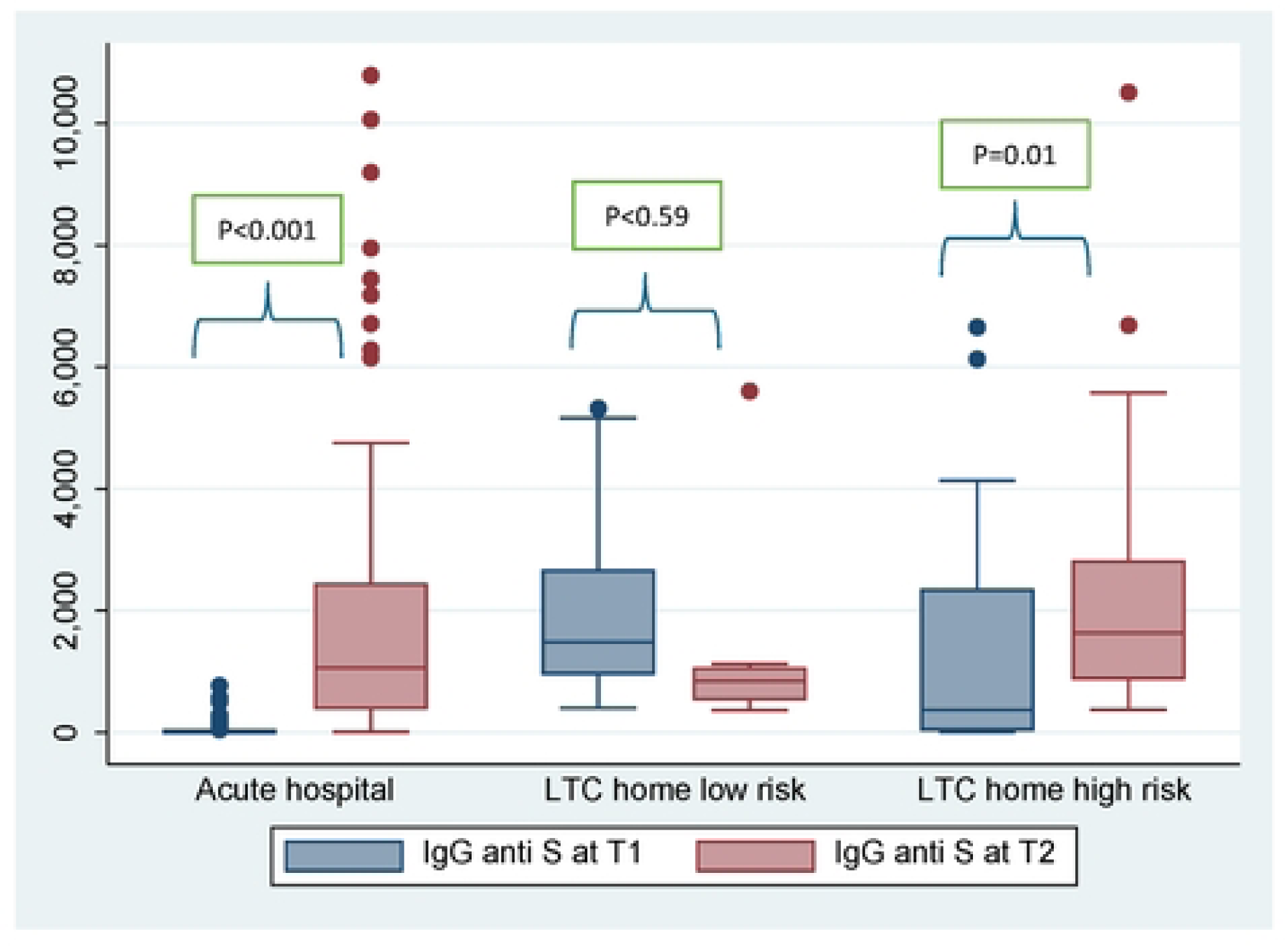
Distribution ofanti-S lgG antibody concentration in HCW at two-time points and by facility type Using non-parametric paired analysis, there was an increase in antibody levels in the acute hospital HCW and in those from the LTC homes at high risk.

### HCW Household and community exposures

Exposure to household COVID-19 cases was also higher during the Omicron wave, especially for HCW in the acute care hospital (Table 6), and at the beginning of the epidemic. Five HCW reported cases of COVID-19 at home pre-Omicron, while 50 HCW reported cases at home during the Omicron wave. The increase in exposure to COVID-19 cases at home during the Omicron wave is consistent with the increase in cases in the community including children [8]. In addition, there was a shift in behaviours othroughoutthe observational period: HCW reported a decreased frequency of use of protective measures during community interactions, specifically reduced use of a mask and social distancing. This behaviour change occurred between the late 2021 and early 2022 periods, data that is consistent with the provincial relaxation of community measures in Ontario in March 2022.

### Vaccination history

By the end of 2022, Health Canada had approved six COVID-19 vaccines but three were predominantly used: Moderna SpikeVax (mRNA, mRNA-1273), Pfizer-BioNTech Comirnaty (mRNA, BNT162b2), and AstraZeneca Vaxzevria (viral vector-based, AZD1222). Vaccination in the cohort started in December 2020. At baseline, 47% of the HCW had been vaccinated, 20% of the acute hospital HCW, 72.7 % of HCW in LTC homes without previous COVID-19 outbreaks, and 36% of the HCW in the LTC homes that had previous COVID-19 outbreaks, reflecting vaccine prioritization HCW and residents of LTC homes [40]. In the acute hospital, HCW received Pfizer for the first, second and the first booster (third dose), except for one person who received AZ as a first dose, 5 who received Moderna and then AZ for their second dose, and 1 who received Moderna initially and then as a booster. In the low-risk LTC homes, most of the HCW received Pfizer except for 3 who received Moderna as booster. In the higher risk LTC homes all received Pfizer except for 1 who had received AZ for first dose; and two who received Moderna as booster. The time interval between first and second doses ranged from 4 to 40 weeks, with a mean of 15 weeks (S2 Figure 8). The mean time between the second and the booster (third dose) was 26 weeks, with a range of 5-78 weeks. In Canada, the approved interval between the second and the booster was ≥ 6 months for immunocompetent individuals (16). In Ontario, the first booster of COVID-19 vaccination started in November 2021 for HCW responding to increasing Omicron cases. At the beginning of the Omicron wave, 15% of the cohort had received one dose, 80% had received two doses, and 3% had received three doses. During the Omicron wave, 65% received a third dose.

### Humoral response

The mean levels at T1 (first blood sample) of IgG against the spike protein were 482 BAU/mL, while at T2 (second blood sample) it was 1913 BAU/mL. At T1, 37.2% had a levels >=35 BAU/mL, while at T2 the proportion at or above this level was 99%. The distribution of IgG levels is presented in Figure 5, at T1, the levels range between 3.2 BAU/mL -6654 BAU/mL, and at T2, between 10.5 BAU/mL -10787 BAU/mL. The higher antibody concentration is consistent with the vaccination status at T2 in all participants. There was no correlation between IgG levels at T1 and T2 (r=0.05). Differences in IgG levels were observed at T1 between institutions (p<0.001) but not at T2 (p=0.07), which likely reflects HCW in the acute care hospitals less likely to be vaccinated before blood assessment at T1 while they had been more exposed to infection and vaccines at T2. (Figure 6). Five participants had positive anti-N antibody titers at T1, 3 of whom reported no previous positive COVID-19 PCR test. At T2, nine HCW were positive, two of whom reported no previous positive COVID-19 PCR test. This suggests that a proportion of individuals had asymptomatic or minimally symptomatic COVID-19 (5 cases) that did not prompt testing, three additional cases before and two after the Omicron wave. Four of the five had IgG levels >35BAU/ml.

## Strengths and weaknesses

Although there are other established cohorts of HCW in Canada during the COVID-19 pandemic, the SCORE study is unique in several ways. This study collects data on factors related to host and the occupational and community environment that influence the risk of acquisition of COVID-19 in HCW. As host factors, the cohort assessed vaccination status, antibody leves, comorbidities and socioeconomic status. This study incorporates detailed surveys of occupational factors, and non-occupational factors. For instance, we inquired about work site, movement within the institution, and the frequency and type of contact with patients and other HCW which allows analyses of mobility and contact variability and risk of COVID-19 [41]. A sizable number of HCW were recruited with a broad spectrum of professions and roles including nurses, physicians, nursing assistants/PSWs, respiratory therapists, and laboratory technicians.Further, our study also quantifies social contact which affects the risk of COVID-19 acquisition. For this latter task, we adapted questions from the Polymod survey to include detailed information on closeness, duration and number of contacts in the community and the household [42]. This analysis will complement previous analysis in Canada and other settings demonstrating the importance of social determinants [43]

The SCORE study needed some adjustments over the months of the epidemic to respond to new knowledge and the rapid deployment of COVID-19 vaccination. This encompassed incorporating the assessment of vaccines such as the number of doses, and the type of vaccines and the type of serologic testing. This resulted in advantages and limitations. For instance, the type of vaccine was not assessed in the first survey as it was not clear when and what vaccines were to be deployed. Once vaccines became available for deployment [44], vaccine type was inquired in the second survey. As a result, at this latter time point, responses were more prone to memory bias and missing information was more frequent for the second vaccine. We attempted to mitigate this limitation by reconstructing probable vaccination dates using the monthly short survey, which offered an acceptable yet modestly imprecise alternative.

Importantly, the number of vaccine doses and the last dose were less likely to be forgotten than the timing. Despite the probable limitations, the obtained descriptive data revealed vaccine timing consistent with the recommended provincial vaccination recommendations.

We are confident that the study captured the vast majority, if not all COVID-19 cases since all HCW with symptoms of COVID-19 were required to be confirmed with PCR testing. It is possible that we missed asymptomatic infections which are reported to be 7.3% or lower [45]. For this, we have assessed the presence of antibodies with Abbot testing, and we found 3 missing infections at baseline and two missing infections during the Omicron wave. These cases will be considered incident COVID-19 cases in future analyses.

This is a convenience sample of HCW and as such it cannot be generalized to HCW working in the same institutions, nor to HCW of LTC homes in the province. However, this cohort resembles the trends in COVID-19 in the communities where the facilities are located, thus describing the experience of a sample of HCW within those settings. Unfortunately, 20% of the sample was lost before the assessment of the second survey possibly biasing the results of the Omicron wave. Loss to follow-up was related to income, smoking and profession, with those at higher risk of COVID-19 being lost to follow-up. Therefore, the end cohort is one with individuals who are likely less at risk of infection compared to the beginning of the cohort.

## Discussion and Planned Analyses

The SCORE cohort offers opportunities to deeply explore the agent, host, and environmental factors influencing COVID-19 transmission in Ontario healthcare workers. In the SCORE cohort, we observed a low incidence of COVID-19 cases until the onset of the Omicron wave, which highlighted the drastic impact on the VOC upon transmission and the importance of infectious agent characteristics. Consistent with other reports in Canadian HCW, the risk of infections in HCW was related to the occupational environment, such as there were a decreased during the beginning of the pandemic due to the use of personal protective equipment and other infection control practices. Remarkably, the acute care hospital was outbreak-free for 16 months and the low-risk LTC homes for at least 24 months from the beginning of the pandemic in Ontario. This was the case even with the emergence of more infectious Alpha and Delta variants in early 2021 with essentially no reported incident cases in the cohort during that time.

Only after the Omicron wave did outbreaks and cases occur in these facilities. Similarly, aspects related to the community environment played an important role in the risk in HCW, that we could futher explore in SCORE. The low prevalence in the region where the cohort was established was likely related to the strict measures in the community, such as closing schools, restrictions in gathering events, mask use and social distancing being reinforced in public settings. An analysis of risk factors for COVID-19 infection with a focus on occupational and community factors is ongoing. Given the distribution of hazards over time, we can focus on the risk in two different periods, at the beginning of the pandemic and during the Omicron wave. Very few studies in HCW have been able to do these longitudinal comparisons [18,46]

Furthermore, qualitative and quantitative assays of humoral response, especially IgG levels to the S protein offer a glimpse into vaccine-vs infection-mediated responses, to establish potential protective levels related to new infection or adverse outcomes. [20] In the SCORE cohort, we will be able to establish the relationship between vaccination status, number of vaccines, and infection and antibody IgG anti-S levels at T1 and T2. Using the Poisson regression as a method to assess associations for highly skewed distributions [47], we will establish the relationships between the number of vaccine doses and previous (yes/no) infection on IgG levels, testing the hypothesis that hybrid immunity confers greater humoral immunity levels. We will also determine whether demographic and health factors are related to higher antibody levels; comparing again levels achieved before and during the Omicron wave.

Furthermore, we have collected information on social contacts in two different ways. First through a weekly diary using the polymod survey, previously used to describe patterns of contacts and to model respiratory disease transmission. And second, through contact tracing analysis of cases. Unfortunately with the high burden of cases and the limited availability of personnel in public health units to do the contact tracing, contact tracing during Omicron was not possible. With the social contact information, we plan a description of the number, risk level, and duration of time spent with social contacts. We will use the social contact survey to identify the type of contacts associated with a higher risk of COVID-19 infection. Egocentric analysis of social contacts will be used to identify differences in social networks in those with and without COVID-19 infection. Networks will also be compared across different settings, acute hospital vs LTC homes.

Data collected in SCORE can further be used to model the effect of interventions in different settings in HCW. For instance, the agent-based model analysis framework proposed by researchers uses data on spatial configuration, use of preventive measures, health and demographics of the individuals, and aspects that are covered by our study [48,49]. The agent-based models can be used to test the reduction of transmission under different future scenarios, testing the model with real data on the movement of HCW in their working and community places.

### Collaboration

The authors are welcoming and encouraging research collaborations using the SCORE data. Data are available on reasonable request and researchers are welcome to contact the research group for further information.

## Data Availability

Data cannot be shared publicly because this has not been approved by our institution. Data are available once a request is done to any of the investigators and access is approved by the Ethics Committee for researchers who meet the criteria for access to confidential data.

## Acknowledgments

We thank the study participants, Dr Gong’s lab team, KS, PZ and BF for their work and assistance in the study collection of data.

## Supporting information

S1 Table 1. The waves of COVID-19 in Canada and SCORE study assessments

S1 Table 2. Descriptive data and sociodemographic characteristics of participants with complete vs incomplete follow-up

S1 Table 3. Table 3s. Reinfections in the cohort of HCW-SCORE 2020-2022.

S2 Figure 1. Provincial Mandates And Institutional Regulations During The Period Covered By The SCORE Cohort

S2 Figure 2: Laboratory confirmed COVID-19 weekly case counts and rates by reported date in region 1, January 12, 2020, to December 31, 2022

S2 Figure 3: Laboratory confirmed COVID-19 weekly case counts and rates by reported date in region 2, January 12, 2020, to December 31, 2022

S2 Figure 4: Laboratory confirmed COVID-19 weekly case counts and rates by reported date in region 3, January 12, 2020, to December 31, 2022

S2 Figure 5. Percent SARS-CoV-2 lineages circulating in Eastern Ontario 12/27/2020 to 08/30/2022

S2 Figure 6. Personal protective equipment used when caring for COVID-19 patients by period and facility type.. Figure 6a corresponds to the proportion of HCW using PPE at beginning of the pandemic, figure 6b depicts PPE during 2021 and figure 6c PPE during Omicron wave.

S2 Figure 7. Personal protective equipment used when caring for COVID-19 patients by period and facility type. Figure 7a corresponds to the beginning of the pandemic, figure 7b, 2021 and figure 7c during the Omicron wave.

S2 Figure 8. Distribution of timing of vaccine doses since the beginning of epidemic (Feb. 1^st^ 2020) in weeks.

